# National hospital readiness for COVID-19 in Lesotho: Evidence for oxygen ecosystem strengthening

**DOI:** 10.1101/2021.04.27.21256199

**Authors:** Jill E. Sanders, Tafadzwa Chakare, Lucy Mapota-Masoabi, Makhoase Ranyali-Otubanjo, Mareitumetse M. Ramokhele, Aleisha Rozario, Eric D. McCollum

**Affiliations:** Jhpiego Lesotho; Ministry of Health, Government of Lesotho; Global Program in Respiratory Sciences, Eudowood Division of Pediatric Respiratory Sciences, Department of Pediatrics, School of Medicine, Johns Hopkins University, Baltimore, Maryland, United States; Department of International Health, Bloomberg School of Public Health, Johns Hopkins University, Baltimore, Maryland, United States

## Abstract

The COVID-19 pandemic continues to challenge healthcare systems globally. To understand hospital capacity, including critical oxygen capacity, a pragmatic assessment of all public hospitals in Lesotho was made in July 2020 (baseline), with follow-up in December 2020. We developed a qualitative hospital services questionnaire modeled on the World Health Organization COVID-19 assessment tool and converted answers into quantitative ordinal variables. At baseline we found all 12 domains had areas demonstrating preparedness and weakness. Key baseline gaps within infection prevention and control were lack of a dedicated team, and insufficient personal protective equipment and space for donning and doffing. Substantial limitations were noted in hypoxemia diagnosis and treatment; information management and care coordination pathways were also suboptimal. Our baseline findings may reflect uneven early pandemic care quality. Targeted follow-up after five months revealed marked improvement in the availability of pulse oximetry, oxygen capacity, and heated high flow nasal cannula devices.

## Introduction

The global pandemic caused by SARS-CoV-2, the novel virus responsible for COVID-19 disease, has impacted health systems at all levels. Sub-Saharan African countries were affected later in the pandemic’s first wave, with peak infections recorded during July 2020^1^. A second wave was experienced during the festive season from December 2020 to January 2021. Lesotho, a nation of two million people in southern Africa, confirmed its first COVID-19 case on May 13, 2020. As of March 3, 2021, a total of 10,521 cases have been confirmed, with 305 deaths^2^.

Lesotho’s health system is comparable to other African countries with a foundation of nurse-led primary healthcare delivered from community-based clinics. Each of Lesotho’s ten administrative districts has at least one secondary-level hospital, which is physician-led and generally lacks higher care. The national tertiary care hospital in Maseru, the capital, is the only government hospital offering intensive care. In response to the pandemic, all district-level hospitals established COVID-19 isolation wards and the Ministry of Health designated two district hospitals as COVID-19 Treatment Centres with inpatient services dedicated to suspected or confirmed cases (Figure 1).

**Figure 1.**
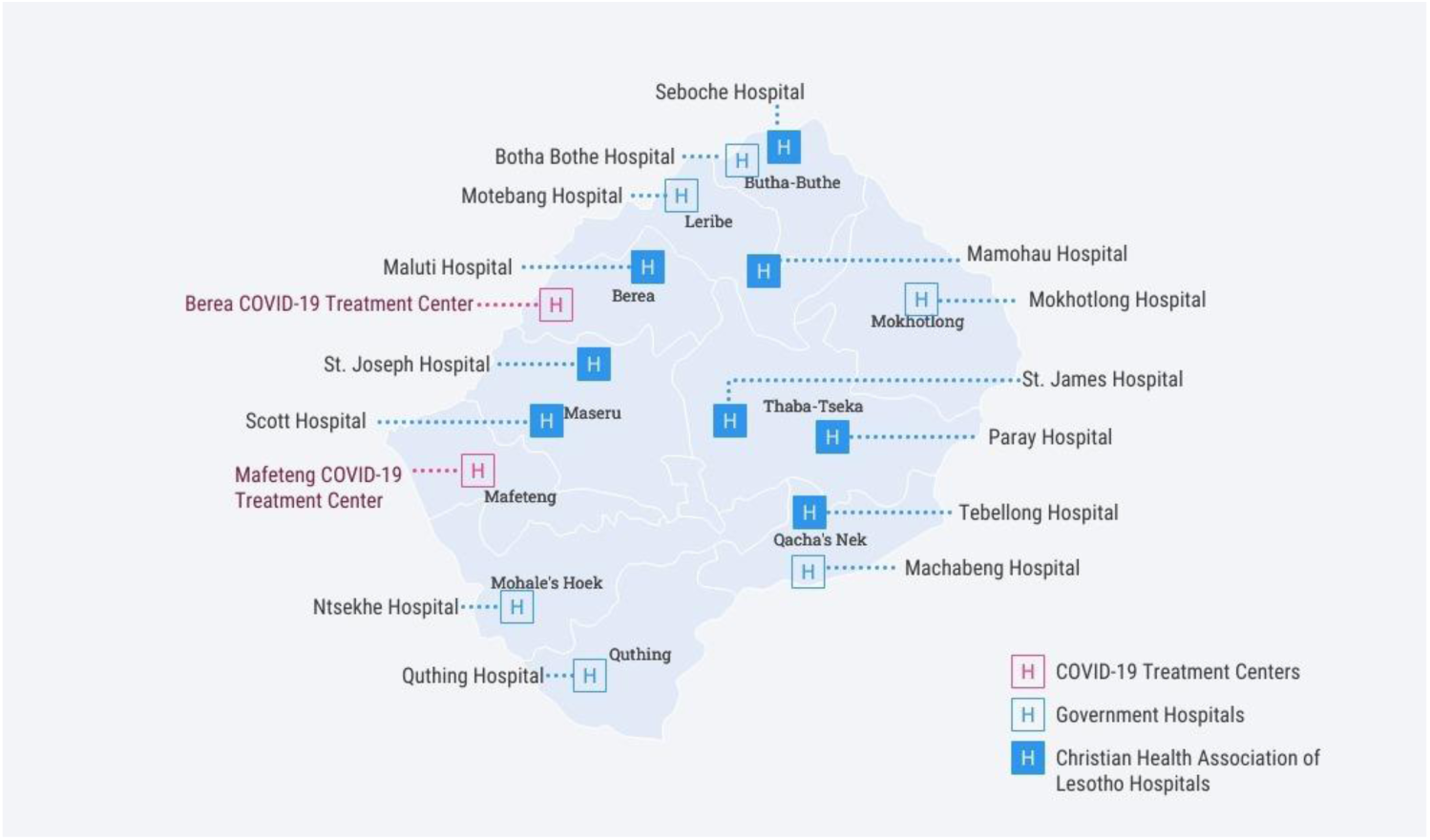
Lesotho map.

There are few publications to date from sub-Saharan African countries on COVID-19 national hospital readiness and oxygen availability^3-6^. To better understand COVID-19 hospital capacity and remaining gaps during the first pandemic wave in July 2020, we conducted a rapid, pragmatic baseline assessment of all public hospitals using an adapted World Health Organization (WHO) COVID-19 hospital assessment tool. We then completed a focused reassessment of the national oxygen ecosystem during the second pandemic wave in December 2020.

## Methods

This is a cross-sectional study of hospital readiness using a researcher-administered questionnaire with additional longitudinal follow-up of the oxygen ecosystem. We developed a questionnaire focused on hospital services to reflect the WHO ‘Rapid Hospital Readiness Checklist’ tool, designed to assist hospital managers in evaluating current and future facility capacities for COVID-19 surges^7^. Questions were grouped into 12 domains: 1) Communication and Coordination, 2) Human Resources Planning, 3) Training, 4) Information Management, 5) Infection Prevention and Control (IPC), 6) IPC Supply Chain Management, 7) Screening, 8) Triage, 9) Diagnostic Testing, 10) Isolation wards, 11) Severe Disease Management, 12) Transport. Assessment options were qualitative and included “Yes or usually”, “Sometimes” and “No or rarely” answers. Answers were converted into quantitative ordinal variables. “Yes or usually” scored one point, “Sometimes” 0.5 points, and “No or rarely” zero points. We supplemented the isolation ward and severe disease management domains with information on the hospital oxygen ecosystem including pulse oximeter, oxygen, and advanced respiratory care availability. We described the distribution of each domain using standard summary statistics and explored post hoc whether there were differences between hospitals by domain using the Kruskal Wallis test. The questionnaire is included as a supplemental appendix. Using The McNemar chi-square test we assessed for any differences in the proportion of hospitals at baseline and follow-up having pulse oximetry at triage and in the isolation ward, and oxygen in the isolation ward. We also analyzed the difference in mean patient-adjusted oxygen capacity per hospital at baseline and follow-up using paired *t*-tests. Patient-adjusted oxygen capacity per hospital was defined as the number of patients that can be simultaneously treated with oxygen per hospital.

Jhpiego Lesotho, a Johns Hopkins University affiliate, is supporting the Lesotho Ministry of Health COVID-19 response to scale-up case management and respiratory care at hospitals with USAID funding. All 16 secondary-level hospitals in Lesotho, including the two COVID-19 Treatment Centres, were visited in person by Jhpiego personnel (JS, MMR) during July 2020 with follow-up in December 2020 to reassess the oxygen ecosystem. Hospital leadership, Medical Superintendents and Managers of Hospital Nursing Services, were the primary responders with support from administrators, human resource managers, pharmacists, and COVID-19 focal persons. When possible, we verified qualitative answers with in-person checks. Constructive feedback was provided to hospital teams.

### Oversight

The survey was conducted in collaboration with Ministry of Health of Lesotho. Consent was not required as Johns Hopkins University Institutional Review Board and Lesotho National Health Research Ethics Committee classified this activity as non-human subjects research.

## Results

### Overall

All 12 surveyed domains had areas demonstrating preparedness and weakness (Table 1). Our post hoc exploratory analysis found no statistical differences between hospitals within any of the 12 domains.

**Table 1.** Lesotho Hospital Readiness Assessment

|  |  | Coordination & Communication (N=7) | HR Planning (N=6) | Training (N=6) | Information Management (N=6) | IPC (N=7) | IPC Supply Chain (N=9) | Screening (N=5) | Triage (N=7) | Diagnostic Testing (N=7) | Isolation Wards (N=5) | Severe Disease Management (N=7) | Transport (N=3) |
| --- | --- | --- | --- | --- | --- | --- | --- | --- | --- | --- | --- | --- | --- |
| District hospitals, % (n) | Scott | 50% (3.5) | 42% (2.5) | 50% (3) | 75% (4.5) | 43% (3) | 39% (3.5) | 70% (3.5) | 86% (6) | 57% (4) | 20% (1) | 86% (6) | 0% (0) |
|  | St Joseph | 79% (5.5) | 58% (3.5) | 58% (3.5) | 67% (4) | 57% (4) | 61% (5.5) | 100% (5) | 50% (3.5) | 29% (2) | 40% (2) | 100% (7) | 0% (0) |
|  | Ntsekhe | 36% (2.5) | 58% (3.5) | 83% (5) | 75% (4.5) | 71% (5) | 100% (9) | 90% (4.5) | 50% (3.5) | 71% (5) | 80% (4) | 86% (6) | 50% (1.5) |
|  | Quthing | 57% (4) | 75% (4.5) | 58% (3.5) | 92% (5.5) | 100% (7) | 56% (5) | 70% (3.5) | 79% (5.5) | 71% (5) | 80% (4) | 71% (5) | 33% (1) |
|  | Machabeng | 100% (7) | 58% (3.5) | 100% (6) | 50% (3) | 64% (4.5) | 89% (8) | 90% (4.5) | 57% (4) | 71% (5) | 80% (4) | 71% (5) | 33% (1) |
|  | Tebellong | 50% (3.5) | 58% (3.5) | 100% (6) | 100% (6) | 50% (3.5) | 78% (7) | 50% (2.5) | 57% (4) | 71% (5) | 20% (1) | 93% (6.5) | 100% (3) |
|  | Paray | 93% (6.5) | 83% (5) | 100% (6) | 100% (6) | 71% (5) | 61% (5.5) | 100% (5) | 36% (2.5) | 71% (5) | 40% (2) | 71% (5) | 50% (1.5) |
|  | St James | 71% (5) | 67% (4) | 100% (6) | 100% (6) | 93% (6.5) | 78% (7) | 80% (4) | 71% (5) | 71% (5) | 70% (3.5) | 79% (5.5) | 33% (1) |
|  | Maluti | 86% (6) | 58% (3.5) | 100% (6) | 58% (3.5) | 71% (5) | 67% (6) | 100% (5) | 64% (4.5) | 71% (5) | 40% (2) | 79% (5.5) | 50% (1) |
|  | Motebang | 43% (3) | 50% (3) | 58% (3.5) | 50% (3) | 57% (4) | 78% (7) | 80% (4) | 71% (5) | 71% (5) | 100% (5) | 79% (5.5) | 33% (1) |
|  | Mamohau | 57% (4) | 75% (4.5) | 67% (4) | 100% (6) | 57% (4) | 94% (8.5) | 90% (4.5) | 100% (7) | 64% (4.5) | 90% (4.5) | 86% (6) | 33% (1) |
|  | Botha Bothe | 100% (7) | 50% (3) | 100% (6) | 58% (3.5) | 64% (4.5) | 67% (6) | 100% (5) | 71% (5) | 71% (5) | 100% (5) | 93% (6.5) | 33% (1) |
|  | Seboche | 57% (4) | 33% (2) | 75% (4.5) | 50% (3) | 64% (4.5) | 72% (5) | 80% (4) | 71% (5) | 71% (5) | 90% (4.5) | 86% (6) | 17% (0.5) |
|  | Mokhotlong | 71% (5) | 67% (4) | 100% (6) | 50% (3) | 86% (6) | 78% (7) | 80% (4) | 43% (3) | 57% (4) | 40% (2) | 100% (7) | 0% (0) |
|  | **Subtotal,**<br>**Mean % (SD)** | **68% (21%)** | **59%**<br>**(13%)** | **82% (20%)** | **73%**<br>**(21%)** | **68%**<br>**(16%)** | **73%**<br>**(16%)** | **84%**<br>**(15%)** | **65%**<br>**(17%)** | **66%**<br>**(12%)** | **64%**<br>**(29%)** | **84%**<br>**(10%)** | **33%**<br>**(26%)** |
| COVID-19<br>Treatment<br>Centre<br>Hospitals,<br>% (n) | Berea | 93% (6.5) | 50% (3) | 100% (6) | 67% (4) | 71% (5) | 83%<br>(7.5) | 90%<br>(4.5) | 64%<br>(4.5) | 71% (5) | 70%<br>(3.5) | 93% (6.5) | 50%<br>(1.5) |
|  | Mafeteng | 93% (6.5) | 42% (2.5) | 100% (6) | 83% (5) | 79% (5.5) | 67%<br>(6) | 90%<br>(4.5) | 86%<br>(6) | 71% (5) | 90%<br>(4.5) | 86% (6) | 50%<br>(1.5) |
|  | **Subtotal,**<br>**Mean % (SD)** | **93% (0%)** | **46%**<br>**(6%)** | **100% (0%)** | **75%**<br>**(11%)** | **75% (6%)** | **75%**<br>**(11%)** | **90%**<br>**(0%)** | **75%**<br>**(16%)** | **71%**<br>**(0%)** | **80%**<br>**(14%)** | **89% (5%)** | **50%**<br>**(0%)** |
| Total | **Mean % (SD)** | **71% (21%)** | **58%**<br>**(13%)** | **84% (20%)** | **73%**<br>**(20%)** | **69%**<br>**(15%)** | **73%**<br>**(15%)** | **85%**<br>**(14%)** | **66%**<br>**(17%)** | **66%**<br>**(11%)** | **66%**<br>**(28%)** | **84% (9%)** | **35%**<br>**(25%)** |

HR indicates human resources; IPC, infection prevention and control; SD, standard deviation.
n=points scored, N=maximum points

### Selected Domains (Table 1)

#### Coordination and communication

Gaps included lack of development and dissemination of internal COVID-19 hospital communications plans.

#### Human Resources Planning

Lack of occupational health plans to address healthcare worker COVID-19 infections and lack of mental health services to staff providing COVID-19 care were consistently observed.

#### Training

Although national COVID-19 Clinical Guidelines were available from 21 April 2020, trainings were still ongoing at 7/16 facilities in July 2020.

#### Information Management

While standardized tools were available for routine collection of clinical and epidemiologic data, there was no clear data management and information flow pathway. Nine facilities had a designated person to collect COVID-19 data, but reporting processes and feedback mechanisms were undefined.

#### Diagnostic testing

No surveyed facility received SARS-CoV-2 testing results directly, requiring facilities to actively pursue results.

#### IPC and IPC Supply Chain Management

Given IPC’s core role in the pandemic response, it was assessed in two sections: first on policies, procedures and infrastructure and second on commodities, including personal protective equipment (PPE). Only 7/16 hospitals had a functioning IPC team. Spaces for staff to safely don (8/16 hospitals) and doff (7/16 hospitals) PPE were limited. All facilities related PPE supply challenges with 8/16 facilities reporting sufficient contact/droplet PPE (gowns, gloves, face shields/goggles) with eye protection supplies limited. Respectively, 9/16 and 7/16 hospitals reported sufficient surgical mask and N95 respirators available for clinical staff. During discussions, facilities volunteered they had adequate PPE only through donations as budgets were insufficient to address high demands. All hospitals also reported challenges ensuring hand hygiene.

#### Screening and Triage

Although all hospitals had COVID-19 screening points for arriving patients, screening procedures like temperature checks and symptom questioning were inconsistently implemented. All 16 facilities had a designated area to provide additional screening and triage to patients reporting a COVID-19 exposure and/or symptoms. However, only three facilities separated symptomatic and asymptomatic persons within triage. Half of hospitals (8/16) reported systematic co-morbidity screening for severe disease among patients presenting with non-severe symptoms.

#### Isolation Wards

COVID-19 isolation wards, designated to serve non-severe patients unable to isolate at home and patients with severe or critical disease awaiting transport to either Treatment Centre, averaged 7 beds (range of zero to 18).

#### Severe Disease Management

The two Treatment Centres care for confirmed or suspected severe or critical COVID-19 cases. Eleven of 14 hospitals reported clinical areas for individuals with severe disease awaiting Treatment Centre transport. Dexamethasone and broad-spectrum antibiotics were available in all hospitals. While chest radiography was possible at 14/16 sites, services were inconsistently available due to concerns around patient flow and IPC.

#### Transport

Transportation of suspect and confirmed COVID-19 patients is coordinated by clinical teams at Treatment Centres, with centralized deployment of ambulances and transport nurses. Non-treatment centre hospitals expressed frustrations with the centralized system, including long delays. Ten facilities reported waiting overnight for ambulances.

#### Oxygen Ecosystem – Baseline in July 2020 (Table 2 and Supplemental Appendix 2)

The diagnosis and oxygen treatment of hypoxemia, a peripheral oxyhemoglobin saturation (SpO_2_) <94%, was limited at all hospitals (Table 2). Pulse oximeters, relatively inexpensive devices used to non-invasively measure SpO_2_, were not available at the entrance of any hospital (i.e., initial screening point), and at symptomatic patient triage (i.e., second screening point) in 10/16 (62.5%) hospitals. Only 9/16 (56.2%) hospitals reported pulse oximeters or vital sign monitors with oximetry functionality at COVID-19 inpatient isolation areas, restricting capacity to monitor patients for new or worsening hypoxemia. While two district hospitals have oxygen plants for medical-grade oxygen production, they provide piped oxygen only to operating theaters and a few patient rooms serving maternity patients. Hospitals, including COVID-19 Treatment Centres, rely upon concentrators and cylinders for oxygen provision. Eleven of 14 hospitals (78.5%) reported access to supplemental oxygen, although the number of patients who could simultaneously receive oxygen varied from one to five, with a total capacity of 28 persons in all non-treatment centre hospitals. Treatment Centres could manage hypoxemia for an additional 22 persons simultaneously, bringing the overall national capacity to 50 patients. Oxygen concentrators with a flow capacity of 5 liters/minute were available in 10/16 hospitals (62.5%). No heated high flow nasal cannula devices or mechanical ventilators were available for COVID-19 patients; only one non-invasive partial pressure ventilation device was identified.

**Table 2.** Lesotho oxygen ecosystem at baseline and follow-up

|  | Pulse oximetry: Triage, % (n/N) |  |  | Pulse oximetry: Inpatient, % (n/N) |  |  | Oxygen, % (n/N) |  |  | Patient-adjusted oxygen capacity per hospital<sup>1</sup>, mean (SD) |  |  |
| --- | --- | --- | --- | --- | --- | --- | --- | --- | --- | --- | --- | --- |
|  | July 2020 | December 2020 | p value<sup>2</sup> | July 2020 | December 2020 | p value<sup>2</sup> | July 2020 | December 2020 | p value<sup>2</sup> | July 2020 | December 2020 | p value<sup>3</sup> |
| District Hospitals | 57.1% (8/14) | 100% (14/14) | 0.0143 | 50.0% (7/14) | 92.9% (13/14) | 0.0143 | 78.5% (11/14) | 100% (14/14) | 0.0833 | 1.9 (1.5) | 6.2 (2.5) | <0.001 |
| COVID-19 Treatment Centres | 100% (2/2) | 100% (2/2) | — | 100% (2/2) | 100% (2/2) | — | 100% (2/2) | 100% (2/2) | — | 11 (4.2) | 30 (0.7) | 0.11 |
| **Total** | 62.5% (10/16) | 100% (16/16) | 0.0143 | 56.3% (9/16) | 93.8% (15/16) | 0.0143 | 81.3% (13/16) | 100% (16/16) | 0.0833 | 3.0 (3.5) | 9.3 (8.6) | <0.001 |

SD indicates standard deviation; COVID-19, coronavirus disease 2019.
<sup>1</sup>Number of patients that can be treated with oxygen simultaneously per hospital
<sup>2</sup>McNemar's Test
<sup>3</sup>Paired *t* test

#### Oxygen Ecosystem – Follow-up in December 2020

Supplies and equipment to diagnose and manage hypoxemia increased after the baseline assessment (Table 2, Supplemental Appendix 3). Follow-up revealed availability of pulse oximeters at all 16 hospitals for symptomatic patient triage, an increase of 37.5% (6/16) over baseline (p=0.0143). Between follow-up and baseline we also observed that 37.5% (6/16) more hospitals made pulse oximeters available within inpatient isolation areas (p=0.0143); only 1/16 (6%) hospitals lacked inpatient pulse oximeters. One of the two district hospitals with an oxygen plant redirected their piped oxygen to supply a five bedded isolation unit. Although cylinders with matched fitted regulators remain scarce, all hospitals had oxygen concentrators. The capacity for oxygen provision increased from 28 to 88 persons (214%) in non-treatment centres and from 22 to 61 persons (177%) in Treatment Centres. Patient-adjusted oxygen capacity increased from an average of 3.0 patients per hospital (SD, 3.5) at baseline to 9.3 patients per hospital (SD, 8.6) at follow-up (p<0.001). Additionally, 11/16 (68.7%) hospitals acquired heated high-flow nasal cannula and 5/16 (31.2%) acquired non-invasive ventilation devices, including both Treatment Centres.

## Discussion

At the time of our baseline assessment of national hospital readiness, Lesotho was in the midst of the peak of its first pandemic wave in July 2020. Our baseline results show inconsistent readiness across all domains interrogated, which likely reflected – to some extent – the quality of COVID-19 hospital care provided during an intense pandemic period. Given the respiratory pathophysiology of SARS-CoV-2, the severe human resource constraints of the Lesotho healthcare system^8^, and the national care pathway designed to centralize COVID-19 hospital resources and expertise at treatment centres, we identified gaps in the hospital oxygen ecosystem as not only the most urgent and feasible to address, but also the most likely to optimize patient outcomes in preparation for subsequent pandemic waves. Our targeted follow-up of the oxygen ecosystem in December 2020 revealed important progress in pulse oximetry availability, oxygen capacity, and the arrival of heated high flow nasal cannula devices and expanded access to non-invasive ventilation for severe COVID-19 patients.

An estimated 19% of symptomatic patients will be hypoxemic, and delivery of supplemental oxygen to patients with severe confirmed or suspected COVID-19 disease is one of the key components to reducing mortality^9,10^. Yet, our baseline July 2020 assessment revealed the limited capacity to diagnose and manage hypoxemia in Lesotho. Most hospitals only had capacity to provide oxygen to one or two patients simultaneously, with the potential for supply to become quickly depleted for extended periods. Our survey did not identify any coordinated systems at the hospital or national level to manage scarce oxygen resources. As Lesotho has stable electricity, concentrators are preferred over cylinders for hypoxemic patients responsive to low oxygen amounts. Cylinders are large, heavy, difficult to transport, and contain finite amounts of oxygen; they are ideally used as a back-up to concentrators. Thus, Lesotho’s relative lack of concentrators and reliance on cylinders necessitated closer monitoring and greater resources to ensure uninterrupted supply. We also observed that even when sufficient quantities of cylinders were present, the regulators required to titrate the cylinder’s oxygen flow were unavailable. Oxygen cylinders cannot be used without an appropriately fitted regulator. Despite notable progress in oxygen availability, we do not know if the observed increase is sufficient to meet future demand. Calculating the estimated burden of severe disease and corresponding oxygen needs is beyond the scope of this analysis.

Neither pulse oximetry nor vital sign monitors with oximetry functionality were available in one-third of Lesotho’s public hospitals at baseline. It has been well established that providers relying only on clinical acumen to prescribe oxygen do so inaccurately, resulting in the misuse of precious oxygen supplies and high proportions of hypoxemic patients failing to receive a potentially life-saving treatment^11,12^. In preparation for possible surges in the COVID-19 pandemic as well as future epidemics, an organized national oxygen system predicated on pulse oximetry and oxygen concentrators was urgently needed in Lesotho.

The Ministry of Health, in collaboration with development partners, was able to rapidly address baseline oxygen equipment shortages for severe COVID-19 disease management. Pulse oximeters were procured and distributed to facilities for use in the diagnosis and monitoring of hypoxemia. Priority was given to procurement of oxygen concentrators, including those with higher flow rates, for provision of supplemental oxygen. An oxygen plant was also completed and opened to support refilling of oxygen cylinders utilized at the Treatment Centres although procurement of cylinder regulators has remained a challenge. Higher levels of respiratory care can now be supported using heated high-flow nasal cannula and non-invasive partial pressure ventilation devices. Effective implementation of both oxygen and higher levels of respiratory care will rely upon ongoing collaboration and human resource capacity building within hospitals and the larger health system.

There are three key limitations to this work. First, this survey was conducted cross-sectionally, and the status of supply chains and stock management are time sensitive. We attempted to account for this through the use of a graded scoring system that recognized partial implementation of interventions and inconsistencies. Second, this survey was based upon self-reporting and given its urgency and our own resource limitations we could not verify all responses. However, our questions were addressed to personnel in the best position to be informed and all respondents were encouraged to consult others. Despite our efforts to optimize reporting accuracy, these results should be interpreted within this context. Lastly, we were unable to follow-up all domains although our baseline assessment identified gaps that would benefit from follow-up.

Delivery of health services in sub-Saharan Africa is complex and the COVID-19 pandemic has placed additional, unparalleled demands upon individuals and systems^13^. Through the use of a user-friendly, pragmatic tool, we conducted a rapid baseline assessment of hospital COVID-19 readiness nationally in Lesotho with targeted follow-up of the oxygen ecosystem. Areas for improvement were noted across all domains. The procurement of pulse oximeters, oxygen concentrators, and advanced respiratory supportive care supplies and equipment, with intensive capacity building of healthcare workers to provide related care is ongoing. Additional support is focused on building capacity and strengthening IPC management and PPE supply chains, along with communications and data sharing. This baseline survey and targeted follow-up served as an initial benchmark, and in anticipation of future viral surges in Lesotho, we recommend repeated surveys to gauge progress and identify persistent gaps across domains in order to optimize patient outcomes.

## Data Availability

The data that support the findings of this study are available from the corresponding author, [JS], upon reasonable request.

## Acknowledgements

We offer our sincere thanks to Ministry of Health and Christian Health Association of Lesotho leadership and personnel for active participation in the survey.

## Author Contributions

Funding acquisition: AR. Conceptualization and design: JS EDM TC AR. Data curation: JS EDM. Data collection: JS. Data analysis: JS EDM. Data interpretation: JS EDM. Writing – original draft: JS. Writing – review & editing: all authors.

## References

1 World Health Organization Coronavirus Disease (COVID-19) Dashboard; https://covid19.who.int, accessed on 2021/03/04.

2 COVID-19 Dashboard by the Center for Systems Science and Engineering (CSSE) at Johns Hopkins University; https://coronavirus.jhu.edu/map.html, accessed on 2021/03/04.

3 Tolu LB, Ezeh A, Feyissa GT. How Prepared Is Africa for the COVID-19 Pandemic Response? The Case of Ethiopia. Risk Manag Healthc Policy. 2020;13:771–776. Published 2020 Jul 9. doi:10.2147/RMHP.S258273

4 Umviligihozo G, Mupfumi L, Sonela N, et al. Sub-Saharan Africa preparedness and response to the COVID-19 pandemic: A perspective of early career African scientists. Wellcome Open Res. 2020;5:163. Published 2020 Sep 16. doi:10.12688/wellcomeopenres.16070.2

5 Nakkazi, Esther. Oxygen supplies and COVID-19 mortality in Africa. The Lancet. Respiratory medicine, S2213-2600(21)00087-4. 10 Feb. 2021, doi:10.1016/S2213-2600(21)00087-4

6 PATH. Biomedical Equipment for COVID-19 Case Management Malawi Facility Survey Report. Seattle: PATH; 2021.

7 World Health Organization Rapid hospital readiness checklist, 26 June 2020. WHO/2019-nCoV/hospital_readiness_checklist/2020.1. Accessed 8 July 2020.

8 The 2018 update, Global Health Workforce Statistics, World Health Organization, Geneva http://www.who.int/hrh/statistics/hwfstats/, accessed on 2021/03/03.

9 Wu Z, McGoogan JM. Characteristics of and Important Lessons From the Coronavirus Disease 2019 (COVID-19) Outbreak in China: Summary of a Report of 72?314 Cases From the Chinese Center for Disease Control and Prevention. JAMA. 2020;323(13):1239–1242. doi:10.1001/jama.2020.2648

10 Jiang B, Wei H. Oxygen therapy strategies and techniques to treat hypoxia in COVID-19 patients. Eur Rev Med Pharmacol Sci. 2020;24(19):10239–10246. doi:10.26355/eurrev_202010_23248

11 McCollum ED, Bjornstad E, Preidis GA, Hosseinipour MC, Lufesi N. Multicenter study of hypoxemia prevalence and quality of oxygen treatment for hospitalized Malawian children. Trans R Soc Trop Med Hyg. 2013;107(5):285–92.

12 Graham HR, Bakare AA, Gray A, Ayede AI, Qazi S, McPake B, et al. Adoption of paediatric and neonatal pulse oximetry by 12 hospitals in Nigeria: a mixed-methods realist evaluation. BMJ Glob Health. 2018;3(3):e000812.

13 Walker PGT, Whittaker C, Watson OJ, et al. The impact of COVID-19 and strategies for mitigation and suppression in low- and middle-income countries. Science. 2020;369(6502): 413–422. doi:10.1126/science.abc0035

