## Supplemental Appendices for "National hospital readiness for COVID-19 in Lesotho: Evidence for oxygen ecosystem strengthening"

**Supplemental Appendix 1: Questions Included in Readiness Questionnaire**

1. Communication and Coordination (7 points maximum)
    a. Are hospital communications systems available and functional? (0, 0.5, or 1 point)
    b. Is the hospital able to communicate with District Response Team regarding COVID-19 prevention and response? (0, 0.5, or 1 point)
    c. Is there a clearly designated person at District Response Team to answer queries? (0, 0.5, or 1 point)
    d. Is there an official hospital spokesperson for COVID-19 information? (0, 0.5, or 1 point)
    e. Is there an internal communications plan for COVID-19 that includes staff responsibilities and contact details? (0, 0.5, or 1 point)
    f. Have hospital staff been briefed on COVID-19 internal communication plans? (0, 0.5, or 1 point)
    g. Are staff regularly briefed on COVID-19 updates to keep everyone well-informed and manage rumors? (0, 0.5, or 1 point)
2. Human Resources Planning (6 points maximum)
    a. Has the hospital administration estimated the current human resources capacity to respond to the potential COVID-19 caseload? (0, 0.5, or 1 point)
    b. Is there a staffing plan to incorporate absenteeism, avoid staff fatigue due to increased workload and ensure continuity of services? (0, 0.5, or 1 point)
    c. Has the hospital's staff directory been updated for use in managing staffing needs during COVID-19? (0, 0.5, or 1 point)
    d. Are there strategies to support reassignment (if necessary) of hospital staff with high risk for complications to manage COVID-19 risks? (0, 0.5, or 1 point)
    e. Is there an occupational health plan to monitor the safety of staff and mitigate COVID-19 risks? (0, 0.5, or 1 point)
    f. Are mental health and psychosocial support services available for staff? (0, 0.5, or 1 point)
3. Training (6 points maximum)
    a. Have clinicians (doctors, midwives, nurses) been trained on COVID-19 Clinical Guidelines to ensure competency and safety? (0, 0.5, or 1 point)
    b. Have other health professionals (pharmacists, social workers, lab technicians) been trained on COVID-19 Clinical Guidelines to ensure competency and safety? (0, 0.5, or 1 point)
    c. Have non-clinical staff (cleaners, kitchen staff, security guards) been trained on COVID-19 Clinical Guidelines to ensure competency and safety? (0, 0.5, or 1 point)
    d. Have administrative staff been trained on COVID-19 Clinical Guidelines to ensure adherence to recommendations? (0, 0.5, or 1 point)
    e. Has District Response Team provided requested support for training? (0, 0.5, or 1 point)

   f. Are COVID-19 Clinical Guidelines available for use by hospital staff? (0, 0.5, or 1 point)
4. Information Management (6 points maximum)
   a. Is there a designated person to ensure the collection and validation of relevant COVID-19 data? (0, 0.5, or 1 point)
   b. Is district COVID-19 Response Team notified for all suspected or confirmed cases, contacts, and travelers? (0, 0.5, or 1 point)
   c. Is DHIS2 (tablet) being used to enter weekly reporting? (0, 0.5, or 1 point)
   d. Are standardized screening tools available? (0, 0.5, or 1 point)
   e. Are standardized laboratory request forms available? (0, 0.5, or 1 point)
   f. Are standardized case identification forms available? (0, 0.5, or 1 point)
5. Infection Prevention and Control (7 points maximum)
   a. Does the hospital have IPC guidelines and standard operating procedures? (0, 0.5, or 1 point)
   b. Does the hospital have a functioning IPC team? (0, 0.5, or 1 point)
   c. Is there signage regarding hand hygiene, respiratory hygiene, and physical distancing posted within the hospital? (0, 0.5, or 1 point)
   d. Are there dedicated areas for staff to safely don (put on) PPE? (0, 0.5, or 1 point)
   e. Are there dedicated areas for staff to safely doff (take off) PPE? (0, 0.5, or 1 point)
   f. Are beds in isolation wards placed at least 1 meter apart? (0, 0.5, or 1 point)
   g. Are beds in general wards placed at least 1 meter apart? (0, 0.5, or 1 point)
6. IPC Supply Chain Management (9 points maximum)
   a. Are cleaning and disinfecting supplies available? (0, 0.5, or 1 point)
   b. Are dedicated cleaning and disinfecting supplies available for isolation areas? (0, 0.5, or 1 point)
   c. Are hand hygiene products (water, soap, hand sanitizer) available for staff? (0, 0.5, or 1 point)
   d. Are hand hygiene products (water, soap, hand sanitizer) available for patients and visitors? (0, 0.5, or 1 point)
   e. Are scrubs available for staff working with COVID-19 cases? (0, 0.5, or 1 point)
   f. Are gowns, gloves, face shields/goggles available for staff requiring contact/droplet precautions? (0, 0.5, or 1 point)
   g. Are surgical masks available for all staff providing clinical care? (0, 0.5, or 1 point)
   h. Are surgical masks available for patients and visitors with respiratory symptoms and/or high-risk COVID-19 exposure? (0, 0.5, or 1 point)
   i. Are N95 masks available for staff performing aerosolizing procedures? (0, 0.5, or 1 point)
7. Screening (5 points maximum)
   a. Are all persons entering hospital grounds (including staff) being screened for COVID-19? (0, 0.5, or 1 point)

b. Is a standardized screening tool being used at all points of entry to the hospital? (0, 0.5, or 1 point)
c. Are all persons performing hand hygiene after screening? (0, 0.5, or 1 point)
d. Are surgical masks being provided for all person with a positive screen? (symptomatic persons, contacts, travelers) (0, 0.5, or 1 point)
e. Have screeners (lay workers and professionals) been trained on screening protocols and case definitions for COVID-19? (suspect case, probable case, confirmed case, contact) (0, 0.5, or 1 point)

8. Triage (7 points maximum)
    a. Is there an area designated for additional screening and triage of those with exposure to COVID-19 or symptoms of COVID-19? (0, 0.5, or 1 point)
    b. Are there separate areas within the triage and isolation area for symptomatic and asymptomatic persons? (0, 0.5, or 1 point)
    c. Is pulse oximetry available in triage to check the oxygen saturation of those with exposure to or symptoms of COVID-19? (0, 0.5, or 1 point)
    d. Are infrared (non-touch) thermometers available for temperature assessment? (0, 0.5, or 1 point)
    e. Are complete vital signs (heart rate, respiratory rate, blood pressure, temperature, oxygen saturation) taken for all persons with exposure to or symptoms of COVID-19? (0, 0.5, or 1 point)
    f. Are those with mild or moderate disease presentation being screened for risk factors for severe disease? (0, 0.5, or 1 point)
    g. Is mental health screening (psychological first aid) provided to all COVID-19 suspect and confirmed cases? (0, 0.5, or 1 point)
9. Diagnostic Testing (7 points maximum)
    a. Are samples for testing being collected on-site? (0, 0.5, or 1 point)
    b. Have all staff collecting samples been trained on specimen collection procedures? (0, 0.5, or 1 point)
    c. Are testing supplies (such as swabs) available? (0, 0.5, or 1 point)
    d. Are you receiving results directly? (0, 0.5, or 1 point)
    e. Are you receiving results within 72 hours? (0, 0.5, or 1 point)
    f. Are specimens transported to National Reference Laboratory within 72 hours? (0, 0.5, or 1 point)
    g. Are diagnostic tests performed for other diagnoses (such as GXP for TB)? (0, 0.5, or 1 point)
10. Isolation Wards (5 points maximum)
    a. Is a log kept of all individuals entering the room of a patient with suspected or confirmed COVID-19? (0, 0.5, or 1 point)
    b. Are designated isolation areas available for asymptomatic contacts or travelers who cannot safely quarantine at home? (0, 0.5, or 1 point)
    c. Are designated isolation areas available for suspect cases with mild/moderate disease who cannot safely isolate at home? (0, 0.5, or 1 point)
    d. Are isolated patients being monitored twice daily for signs of deterioration? (0, 0.5, or 1 point)

    e. Is pulse oximetry available in isolation areas to monitor for deterioration? (0, 0.5, or 1 point)

11. Severe Disease Management (7 points maximum)
    a. Are designated isolation areas available for suspected or confirmed cases with severe disease awaiting transport to Treatment Centre? (0, 0.5, or 1 point)
    b. Is supplemental oxygen available for patients with severe disease awaiting transport to Treatment Centre? (0, 0.5, or 1 point)
    c. Are nasal cannulas and face masks available for patients need supplemental oxygen? (0, 0.5, or 1 point)
    d. Is dexamethasone available for patients with severe disease? (0, 0.5, or 1 point)
    e. Are broad-spectrum antibiotics available for patients with severe disease? (ceftriaxone, azithromycin, clarithromycin, amoxicillin-clavulanate, doxycycline) (0, 0.5, or 1 point)
    f. Is radiology (CXR) available for patients with severe disease? (0, 0.5, or 1 point)
    g. Is rapid glucometer available to monitor blood sugar? (0, 0.5, or 1 point)
12. Transport (3 points maximum)
    a. Is the ambulance available on time? (0, 0.5, or 1 point)
    b. Is District Health Management Team able to follow-up alerts? (0, 0.5, or 1 point)
    c. Is transportation available for contact tracing and other community outreaches? (travelers, suspect cases, etc.) (0, 0.5, or 1 point)

**Supplemental Appendix 2:** Lesotho Oxygen Ecosystem at Baseline in July 2020

| | | Pulse oximetry: Triage (yes/no) | Pulse oximetry: Inpatient (yes/no) | Oxygen (yes/no) | Oxygen cylinders with regulators, N | Oxygen concentrators, N | Oxygen delivery capacity, N | Heated high flow nasal cannula (yes/no) | Non-invasive partial pressure ventilation (yes/no) | Invasive mechanical ventilation (yes/no) |
|---|---|---|---|---|---|---|---|---|---|---|
| District Hospitals | Scott | Yes | No | Yes | 0 | 1 | 1 | No | No | No |
| | St Joseph | No | Yes | Yes | 0 | 4 | 4 | No | No | No |
| | Ntsekhe | No | No | No | 0 | 0 | 0 | No | No | No |
| | Quthing | Yes | Yes | No | 0 | 0 | 0 | No | No | No |
| | Machabeng | No | No | No | 0 | 0 | 0 | No | No | No |
| | Tebellong | Yes | No | Yes | 1 | 1 | 2 | No | No | No |
| | Paray | No | No | Yes | 3 | 2 | 5 | No | No | No |
| | St James | No | No | Yes | 3 | 0 | 3 | No | No | No |
| | Maluti | Yes | Yes | Yes | 0 | 3 | 3 | No | No | No |
| | Motebang | Yes | Yes | Yes | 1 | 0 | 1 | No | No | No |
| | Mamohau | Yes | Yes | Yes | 1 | 0 | 1 | No | No | No |
| | Botha Bothe | Yes | Yes | Yes | 0 | 2 | 2 | No | No | No |
| | Seboche | Yes | Yes | Yes | 0 | 3 | 3 | No | No | No |
| | Mokhotlong | No | No | Yes | 0 | 2 | 2 | No | No | No |
| | Sub-Total, % (n/N) | 57.1% (8/14) | 50.0% (7/14) | 78.5% (11/14) | 10 | 18 | 28 | 0% (0/14) | 0% (0/14) | 0% (0/14) |
| COVID-19 Treatment Centre Hospitals | Berea | Yes | Yes | Yes | 6 | 8 | 14 | No | Yes (1) | No |
| | Mafeteng | Yes | Yes | Yes | 6 | 2 | 8 | No | No | No |
| | Sub-Total, % (n/N) | 100% (2/2) | 100% (2/2) | 100% (2/2) | 12 | 10 | 22 | 0% (0/2) | 50.0% (1/2) | 0% (0/2) |
| **Total** | | 62.5% (10/16) | 56.3% (9/16) | 81.3% (13/16) | 22 | 28 | 50 | 0% (0/16) | 6.2% (1/16) | 0% (0/16) |

Supplemental Appendix 3: Lesotho Oxygen Ecosystem at Follow-up in December 2020

| | | Pulse oximetry: Triage (yes/no) | Pulse oximetry: Inpatient (yes/no) | Oxygen (yes/no) | Oxygen cylinders with regulators, N | Oxygen concentrators, N | Oxygen delivery capacity, N | Heated high flow nasal cannula (yes/no) | Non-invasive partial pressure ventilation (yes/no) | Invasive mechanical ventilation (yes/no) |
|---|---|---|---|---|---|---|---|---|---|---|
| District-based Hospitals | Scott | Yes | Yes | Yes | 1 | 3 | 4 | Yes | Yes | No |
| | St Joseph | Yes | Yes | Yes | 2 | 4 | 6 | No | No | No |
| | Ntsekhe | Yes | Yes | Yes | 2 | 6 | 8 | Yes | No | No |
| | Quthing | Yes | Yes | Yes | 0 | 4 | 4 | Yes | No | No |
| | Machabeng | Yes | Yes | Yes | 1 | 6 | 7 | Yes | No | No |
| | Tebellong | Yes | No | Yes | 1 | 2 | 3 | No | No | No |
| | Paray | Yes | Yes | Yes | 3 | 10 | 13 | No | Yes | No |
| | St James | Yes | Yes | Yes | 3 | 3 | 6 | No | No | No |
| | Maluti | Yes | Yes | Yes | 5 | 3 | 8 | Yes | Yes | No |
| | Motebang | Yes | Yes | Yes | 1 | 7 | 8 | Yes | No | No |
| | Mamohau | Yes | Yes | Yes | 1 | 2 | 3 | No | No | No |
| | Botha Bothe | Yes | Yes | Yes | 0 | 6 | 6 | Yes | No | No |
| | Seboche | Yes | Yes | Yes | 0 | 6 | 6 | Yes | No | No |
| | Mokhotlong | Yes | Yes | Yes | 0 | 6 | 6 | Yes | No | No |
| | Sub-Total, % (n/N) | 100% (14/14) | 92.9% (13/14) | 100% (14/14) | 20 | 68 | 88 | 64.3% (9/14) | 21.4% (3/14) | 0% (0/14) |
| COVID-19 Treatment Centers | Berea | Yes | Yes | Yes | 9 | 21 | 30 | Yes | Yes | No |
| | Mafeteng | Yes | Yes | Yes | 10 | 21 | 31 | Yes | Yes | No |
| | Sub-Total, % (n/N) | 100% (2/2) | 100% (2/2) | 100% (2/2) | 19 | 42 | 61 | 100% (2/2) | 100% (2/2) | 0% (0/2) |
| **Total** | | 100% (16/16) | 93.8% (15/16) | 100% (16/16) | 39 | 110 | 149 | 68.8% (11/16) | 31.2% (5/16) | 0% (0/16) |
